# Effectiveness of a physical therapy program based on therapeutic physical exercise and health education to improve quality of life and health status in major depressive disorder: A mixed-method pilot study protocol

**DOI:** 10.1101/2025.05.25.25328327

**Authors:** José Lesmes Poveda-López, Carolina Jiménez-Sánchez, Raquel Lafuente-Ureta, Bárbara Gómez-Marco, Ana Villagrasa-Cantín, Sara Pérez-Mansilla, Marta Guarch-Rubio, Juan Francisco Roy-Delgado

**Affiliations:** Department of Physical Therapy, Faculty of Health Sciences, Universidad San Jorge, Villanueva de Gállego, Zaragoza, Spain; Department of Psychiatry, Royo Villanova Hospital, Health Service of Aragón, Universidad de Zaragoza, Zaragoza, Spain; Department of Psychology, Faculty of Health Sciences, Universidad San Jorge, Villanueva de Gállego, Zaragoza, Spain; Faculty of Health Sciences, Universidad Internacional de La Rioja

**Keywords:** Major Depressive Disorder, Physical Therapy Modalities, Exercise Therapy, Health Education, Quality of Life, Hospitals

## Abstract

**Background:** Major depressive disorder is a mood disorder with significant psychological and physical symptoms, leading to disability and severe consequences. It is influenced by genetic and environmental factors, causing neurotransmitter imbalances and inflammation. Given the high prevalence and impact, it is crucial to implement a health promotion and intervention program. Investigating the efficacy of physical therapy, including therapeutic exercise and health education, compared to psychiatric and psychological approaches, is essential to improve the quality of life for these patients.

**Methods:** A concurrent nested mixed-methods study with quantitative dominance will be conducted. The quantitative study will be a quasi-experimental pilot study with a pre-post design. Additionally, this study includes a qualitative narrative design. Initial and post-intervention evaluations will include sociodemographic and clinical data. Quantitative data will be collected using the EQ-5D-3L, MADRS, NRS, GSE, and GCPC-UN-ESU questionnaires. These tools assess health status, depression severity, pain intensity, self-efficacy, and satisfaction levels. Qualitative data will be collected from focus groups with 6-8 participants. The question guide for patients will cover experiences with illness and intervention, while the guide for professionals will cover perceptions of patient management and observed barriers and facilitators. All participants will receive the same evidence-based intervention over 3 to 6 weeks, with 2 weekly sessions of approximately 45 minutes each. Each session will consist of Therapeutic Exercise and Health Education to improve physical condition and self-management skills.

**Discussion:** This study aims to fill the knowledge gap on the effectiveness of physical therapy interventions for quality of life, pain, and self-efficacy in hospitalized patients with major depressive disorder. It will evaluate the impact of therapeutic exercise and health education, alongside standard psychiatric, psychological, and nursing treatments. The findings will provide scientific insights and guide healthcare policy makers to incorporate physical therapy into hospital treatments for major depressive disorder.

**Clinical Trial registration:** NCT06983405

## Introduction

The World Health Organization[1] indicates that mental health is the state of health that enables individuals to cope with stress through their abilities and skills, and that it is a fundamental human right and a necessary element for community development. Its decline leads to mental illness, characterized by distress, functional disability, increased difficulty in basic and instrumental activities of daily living, sedentary lifestyle, muscle atrophy, poor quality of life, low muscle strength and fatigue, pain, changes in muscle tone, cognitive and affective deterioration, social isolation, stigmatization, work absenteeism, risk of self-harm and premature death. Additionally, it generates an increased burden on health services, resulting in higher costs[2].

According to the International Classification of Diseases, Tenth Revision (ICD-10)[3] and the Diagnostic and Statistical Manual of Mental Disorders, Fifth Edition (DSM-5) by the American Psychiatric Association[4], major depressive disorder (MDD) is a mood disorder characterized by profound sadness and a sustained loss of interest in any activity for at least two weeks. Clinical depression is associated with a significant loss of quality of life, accompanied by psychological symptoms [anhedonia, anergia, sleep and appetite disturbances] and physical symptoms due to functional impairment. It is the most common cause of disability worldwide[5], affecting women[6] and individuals with lower economic income[7,8] more severely. In addition, it causes severe suffering and disruption to work, family, and social activities, with the most severe consequence being suicide[9,10]. Its clinical manifestation may include a single episode or recurrent ones over more than two years, ranging from mild-moderate to severe[11]. All these issues have a significant impact on the population and require a comprehensive approach to their management[12].

MDD is a multifactorial disease that is influenced by genetic and environmental factors[13]. The presence of these factors leads to a decrease in the neurotransmitters: serotonin, norepinephrine, and dopamine. This dysfunction is mediated by hyperactivity of the hypothalamic-pituitary-adrenal (HPA) axis, which is responsible for stress control and the cortisol presence, and a decrease in the morphological volume of the hippocampus due to reduced dendrites and neuronal networks[14,15]. MDD is considered an inflammatory disease due to the increased levels of pro-inflammatory cytokines in the blood of these patients, caused by the activation of the HPA axis[5,16].

The prevalence of mental health disorders is 17%[17] in Europe and reaches 27.4% in Spain[1]. MDD is the mental illness with the highest incidence rate in the adult population, affecting 6.4% of the European population and 4.1% of the Spanish population[18]. In the first year after the COVID-19 pandemic, the prevalence of mental illnesses, especially depressive and anxiety disorders, increased by 25% worldwide[19].

The aging process of the population must also be considered, as currently, 19% of the population in Spain is over 65 years old, and in 10 years, it is expected to be more than 25%. In this population, poor mental health status leads to a higher risk of comorbidity and disability[20], loneliness, stigmatization, and institutionalization[21,22], which contribute to an increased risk of mortality[23,24]. The conditions of aging require closer monitoring due to the severity of their repercussions and the increase in healthcare costs and resource consumption[25].

The usual treatments for MDD are multimodal and include primary and specialized medical and psychological care[26–28], usually combined with pharmacological[29–31] treatment, and to a lesser extent, with physical therapy[32,33]. Although pharmacology can be effective, lack of adherence and adverse effects can compromise the patient’s autonomy[34]. Psychotherapy, including techniques such as cognitive-behavioral therapy, is a common therapy for treating the affective and emotional symptoms of MDD[35,36]. Regarding physical therapy, physical therapists deal with movement problems related to age, injury, pain, or illness, addressing both physical and psychological, emotional, and social aspects in order to prevent, maintain, and restore movement and functional capacity throughout life. Physical therapy must focus on the needs of the population and work in multimodal teams to improve the mental health of patients while always adhering to ethical principles[37]. Thus, physical therapy can enhance the body-mind connection through health education[38] and movement therapy, increasing the patient’s awareness of their capabilities and needs[39]. It addresses physical symptoms such as pain, strengthens the patient[40–42], and improves their mood and functionality through the release of endorphins[43]. Therapeutic exercise ensures professional and coordinated treatment that enhances the patient’s emotional and physical condition[33].

Given the high prevalence and severe impact of MDD on adult patients’ health, the community, and the healthcare system, it is crucial to implement a multimodal coordinated program of health promotion, intervention, and education. The WHO emphasizes the importance of mental health and recommends action plans that prioritize mental health, taking into account physical, social, and economic conditions, and strengthen community support networks. Therefore, it is appropriate to investigate the efficacy of physical therapy treatments, including health education and therapeutic exercise, compared to exclusively psychiatric and/or psychological approaches, to improve the quality of life and health status of patients with MDD in short-stay psychiatric units.

Our study aims to 1) to analyze the efficacy of a physical therapy intervention program based on health education and therapeutic exercise for adults with MDD in a short-stay psychiatric unit, and 2) to explore the perceptions of disease management, barriers and facilitators related to the multimodal treatment among the study participants and the healthcare professionals of the psychiatric team.

## Materials and Methods

### Design

A concurrent nested mixed-methods study with quantitative dominance will be conducted. The quantitative study will be a quasi-experimental pilot study with a pre-post design. Additionally, this study includes a qualitative narrative design, in which focus groups will be conducted to analyze, through content analysis, the experience of the intervention, as well as the perceptions regarding the management of their process, and the barriers and facilitators encountered by the participants during the implementation of the intervention program. Furthermore, the perception of the professionals [both healthcare and non-healthcare] from the Psychiatry Department at Royo Villanova Hospital in Zaragoza regarding the proposed intervention program, and how they perceive it has influenced the disease management, barriers and facilitators, for the patients admitted to their unit will be investigated.

This study will be divided into two phases. Phase 1 will focus on the development of quantitative design, and Phase 2 will include the development of qualitative design through focus groups (Fig 1).

**Fig 1.**
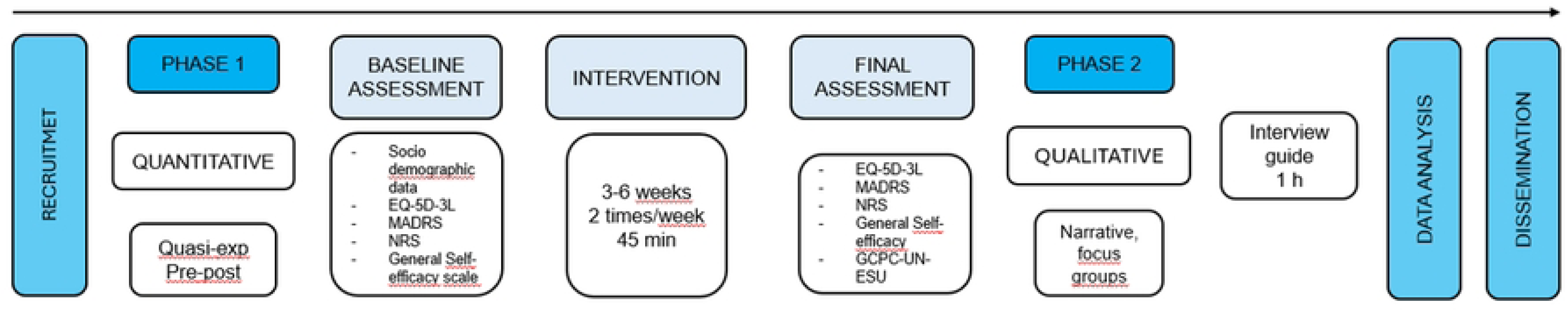
Overall Project Timeline and Phases.

### Ethical approval and considerations

The study protocol was approved by the San Jorge University Ethic Committee (N° 38/3/24-25) and Aragon Ethic Committee (PI25-233).

All registrations and focus group discussions will take place in private locations. Names will not be associated with notes or other study materials. Free and informed consent will be obtained from participants. Data will be documented electronically on data collection tablets. A separately completed declaration of consent will be required for all recordings made during the interviews. As part of the informed consent procedure, participants are informed that the anonymized data may be used for publication.

The study will be conducted in accordance with the Declaration of Helsinki and following the guidelines of the Mixed Method Reporting in Rehabilitation & Health Sciences (MMR-RHS)[44] (S1 Appendix I).

### Participants and eligibility criteria

Two different groups will be involved in the project: patients admitted to the short-stay unit of the Psychiatry Department at Royo Villanova Hospital in Zaragoza, who will participate in both phases of the study (quantitative and qualitative); and the professionals from this department, who will only participate in phase 2 (qualitative).

Thus, the sample will consist of consecutive voluntary patients admitted to the short-stay unit of the Psychiatry Department at Royo Villanova Hospital (Zaragoza) who meet the following inclusion criteria: over 18 years of age; admitted to the psychiatry unit of Royo Villanova Hospital; diagnosed by a physician with MDD; under regular medical, psychological, and pharmacological treatment for their illness; and no need for supervision and control by professional staff of the psychiatric unit during data collection and intervention. Exclusion criteria: patients with comorbid physical or mental illness whose clinical characteristics and/or severity prevent understanding and/or following physical therapy interventions, presence of physical or mental dysfunction or disability that constitute a total or partial contraindication for physical therapy techniques, legal incapacity or pregnancy. Finally, dropout criteria will be considered if the subject’s will express desire to withdraw from the study, if attendance of less than 80% of the physical therapy intervention sessions will be attended, if an injury prevents the continuation of the sessions or if a new disease occurs whose diagnosis and/or severity prevents the continuation of the study or results in death.

Phase 2 will incorporate an additional sample of eight individuals who will participate in a focus group for the qualitative evaluation of professionals comprising the Psychiatry professional team, including both healthcare and non-healthcare staff, at the Hospital Royo Villanova in Zaragoza. Participants for the focus group will be recruited through purposive sampling. Participation will be voluntary, and individuals must meet the inclusion criterion of being healthcare or non-healthcare professionals from the short-stay Psychiatry unit of the Hospital Royo Villanova in Zaragoza. Exclusion criteria include professionals from the unit who were not actively employed under a single, permanent, or temporary employment contract throughout the duration of the physical therapy intervention.

Withdrawal criteria encompass the explicit desire of a participant to withdraw from the study, sick leave or death, or job transfer to another unit or healthcare center.

### Sample size

A sample size of 50 subjects will be required for phase 1 of the pilot study as it ensures manageability, allows for the detection of preliminary effects, conserves resources, provides adequate variability, and is consistent with scientific precedent[45,46] (Fig 2).

**Fig 2.**
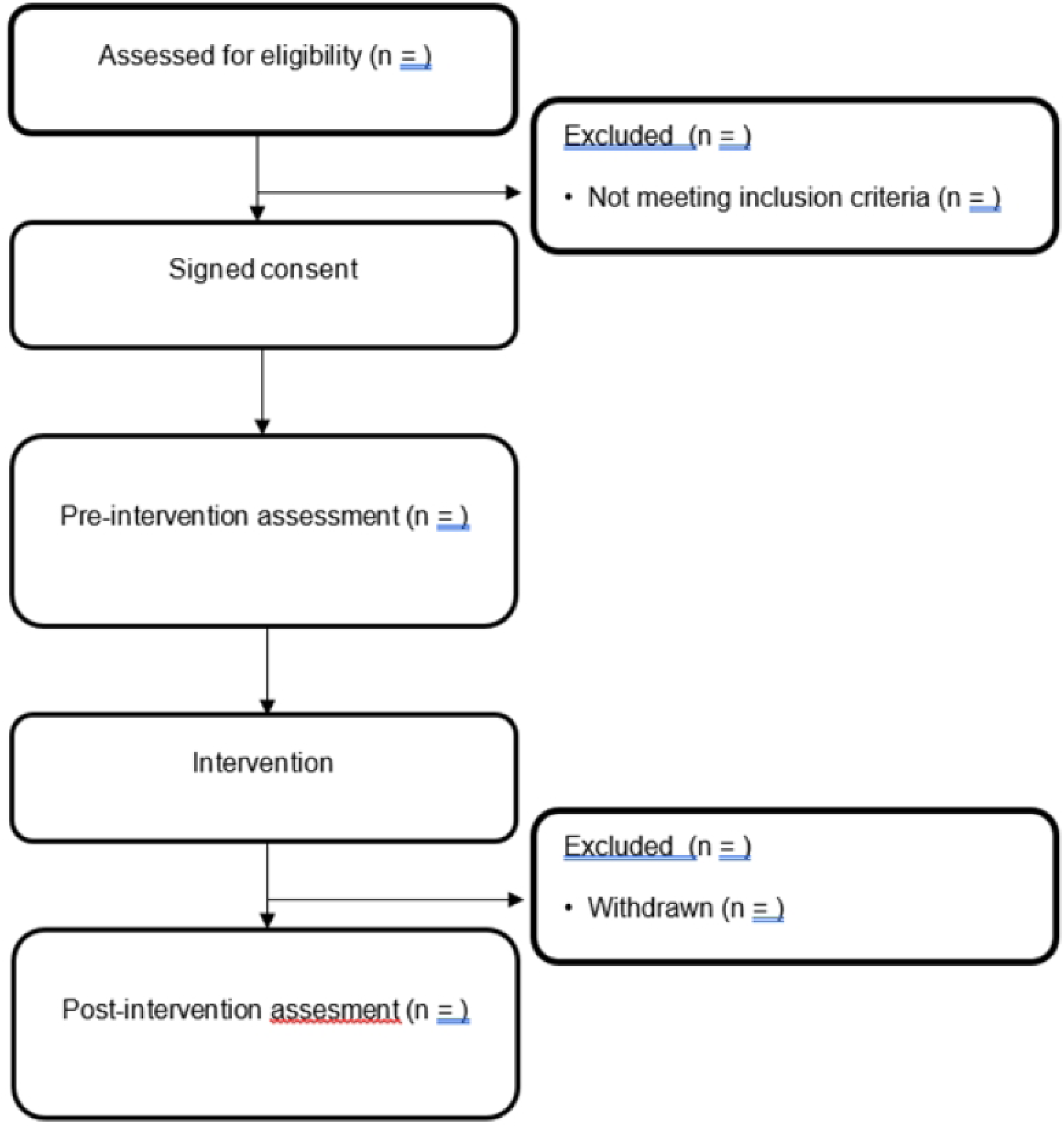
Flowchart.

For phase 2, a purposeful sampling will be led, including MMD patients and different focus groups (for patients and professionals) with a sample of 6-8 participants in each one will be needed until data saturation will be reached[47,48].

### Procedure

#### Quantitative data collection

An initial assessment will always be carried out before the start of the procedure and another at the end of the procedure before discharge from hospital. The initial assessment will include measurements such as age, gender, occupation and clinical data (height, weight, body mass index, medical diagnosis, pharmacological treatment and other non-pharmacological treatments).

Quantitative data will be collected using the following questionnaires and scales: the EuroQol-5D-3L Spanish version questionnaire (EQ-5D-3L)[49], the Montgomery-Asberg Depression Rating Scale (MADRS) Spanish version[50], the Numerical Rating Scale (NRS)[51], the General Self-Efficacy Scale (GSE) Spanish version[52] and the Health Care Satisfaction Survey for People with Chronic Illness (GCPC-UN-ESU)[53].

The EQ-5D-3L health questionnaire can be self-administered or conducted via interview, assessing the patient’s health status across various dimensions (mobility, pain, mental health, etc.), consists of 5 questions with 3 response options (good health=1, some problems=2, or health problems=3), where responses are coded and coefficients applied to reach a reference value and it includes a 20 cm vertical visual analog scale, with values from 0=worst health state to 100=best health state, where the patient indicates their current perceived health status.

The MADRS is used for detecting and assessing the severity of major depressive disorder and consists of 10 questions about cognitive and affective/emotional symptoms, with response values ranging from 0 to 6 (0=least symptomatic and 6=most symptomatic) and its interpretation is: 0-6 points no depression, 7-19 points mild depression, 20-34 points moderate depression, and 35-60 points severe depression.

The GSE consists of a 10-question questionnaire that measures an individual’s perception of their ability to manage life in stressful situations, and each question is answered on a Likert scale from 1 (totally disagree) to 5 (totally agree), being it scores between 27 and 38 points considered average self-efficacy.

Finally, the GCPC-UN-ESU is a validated tool for measuring satisfaction levels that contains 19 items across 4 dimensions: care, health education, service quality, and service loyalty and each item is scored from 1 (not satisfied) to 5 (extremely satisfied); higher scores indicate greater satisfaction, and this will be measured only in the post-intervention evaluation and anonymously.

#### Qualitative data collection

Qualitative data will be collected from two different population groups (MDD patients and professionals) using focus groups with 7-8 participants, several focus groups will be conducted until data saturation. All focus groups will be led by one researcher, while another researcher will take field notes and impressions from participants during the focus group. Each focus group will last approximately 1 hour, based on a question guide, and will be recorded with a recorder after obtaining signed consent from each participant. The question guide in MMD patients’ group will cover experiences with illness, perceptions of process management, and barriers and facilitators encountered during the intervention program. The question guide in professionals’ groups will cover perceptions of patient disease management and the barriers and facilitators observed during the intervention program (S2 File).

### Intervention

This intervention is grounded in the growing body of evidence supporting the antidepressant effects of exercise[54,55] and the importance of health education in promoting self-management and long-term adherence[56].

The therapeutic exercise component will consist of 45-minute sessions, conducted twice weekly, over a period of 3 to 6 weeks[57], adjusted to the participant’s length of hospital stay. Each session will include: 1) Active joint mobility: Involving gentle and controlled exercises to improve or maintain the range of motion in major joints. 2) Strength exercises: Using body weight and/or elastic bands, involving low-to-moderate intensity exercises utilizing the participant’s own body weight and/or progressive resistance elastic bands, targeting major muscle groups. This is included as strength training has demonstrated moderate effects in reducing depressive symptoms[58] and may contribute to improving overall physical health in individuals with mood disorders[59]. 3) Balance exercises: To enhance stability and prevent falls, with difficulty adjusted to each participant’s level. 4) Progressive muscle relaxation: Implemented at the end of each session to reduce muscle tension and promote relaxation[60].

This exercise program will be designed to be progressive and adaptable to the individual capabilities of the patients, taking into account their clinical status and potential physical limitations. Active participation and modification of exercises as needed will be encouraged.

The health education component will be integrated into the exercise sessions and may include brief discussions and informational materials on the following topics: 1) The relationship between physical activity and mental health, explaining potential biological and psychological mechanisms through which exercise can ameliorate depressive symptoms[61]. 2) The benefits of the specific therapeutic exercises included in the program (strength, balance, mobility) concerning improvements in mood, physical functioning, and quality of life[62]. 3) Strategies for increasing daily physical activity after hospital discharge, promoting long-term adherence[58] and the integration of exercise as a regular lifestyle component. 4) Self-assessment and monitoring techniques for physical activity and mood to foster self-efficacy and self-management[58]. 5) Information on community resources and physical activity programs available post-discharge[58].

The intervention will be delivered by professionals trained and experienced in therapeutic exercise and mental health. Adherence to exercise sessions will be monitored, and any potential adverse events will be recorded. Patient response will be evaluated using depression and physical functioning rating scales, both at baseline and at the end of the intervention (S3 table).

### Data analysis

#### Quantitative Data Analysis

The statistical analysis of the quantitative data will be performed using IBM-SPSS Statistics version 29.

Data will be expressed as mean and standard deviation or median and interquartile range. The Shapiro-Wilk test will be used to assess normality. Simple and/or repeated measures ANOVA with post hoc tests will be conducted to compare pre- and post-test scores. If the assumptions for ANOVA are not met, non-parametric tests with post hoc tests will be used. The significance level will be defined as p ≤ 0.05.

Effect size will be calculated using Cohen’s d to determine clinical significance: insignificant, small, medium, and large differences will be reflected in effect sizes of <0.2, 0.2-0.5, 0.5-0.8, and >0.8, respectively[63].

#### Qualitative Data Analysis

The qualitative data obtained from the focus groups will be analyzed using content analysis. Sessions will be fully transcribed verbatim by a single researcher from the recordings made during the focus groups. Initially, one or more complete readings will be conducted to obtain a global understanding of the recorded information and achieve immersion in the text. Subsequently, a second word-by-word reading will be performed, during which inductive coding of the transcripts will be carried out to capture key concepts and thoughts: meaning units will be transformed into condensed meaning units, then into codes[64]. The codes will then be grouped by their relationship and linkage into subcategories. Depending on the relationships between subcategories, the researchers may compare, combine, or organize this larger number of subcategories into a smaller number of categories and main categories[65]. This process will be conducted independently by two researchers[66].

The research team will discuss the categories and subcategories derived from the related narrations, refining them until reaching a consensus. Trustworthiness of the data will be ensured through data and investigator triangulation[67]. Additionally, an integration of quantitative and qualitative data will be conducted.

### Consideration of Gender Perspective

Equitable inclusion across different sexes will be taken into account during recruitment to achieve a point prevalence per sex similar to the latest available data from the health sector or the general population in the city of Zaragoza or the Autonomous Community of Aragon[68]. Major depressive disorder is a disease with a higher prevalence in the female sex[69], therefore, consideration will be given to whether there are differences according to sex in order to offer specific solutions.

## Discussion

Despite the growing interest in physiotherapy interventions, there remains a significant gap in understanding their impact on quality of life, pain management, and self-efficacy among individuals with MDD, particularly during the acute phase and hospitalization. This study will significantly contribute to expanding the knowledge about the effects that a physiotherapy intervention program based on therapeutic exercise and health education can offer this to vulnerable population, as a complement to standard psychiatric, psychological, and nursing treatment.

The findings of this research will not only provide relevant scientific evidence but also valuable information to guide health policy makers in the potential implementation of physiotherapy as an integral part of the multidisciplinary approach to MDD in the hospital setting.

The marked prevalence and profound impact of MDD on the health of adult patients, in the community, and in the health system overall, underscore the urgent need to explore alternative, rapid, and easily accessible treatment options[70]. Depressive illness, in its progression, can intensify and/or trigger the development of physical[33] and mental[71] comorbidities, in addition to impacting a reduced life expectancy[72].

The choice of a mixed-methods design with quantitative predominance, of a nested mixed-methods design, is based on the need to obtain a comprehensive understanding of the intervention’s impact[73]. The quantitative phase, through a pre-post pilot quasi-experimental design, will enable the evaluation of changes in quality of life, pain, self-efficacy, depression severity, and satisfaction with care. Concurrently, the qualitative phase will use focus groups with patients and healthcare professionals to examine in detail their experiences with the intervention, their perceptions of disease management, and the factors that act as barriers or facilitators in the implementation of the program. This integration of quantitative and qualitative data will enable the triangulation of findings[74], providing a richer and more contextualized understanding of the studied phenomenon, surpassing the insights offered by a unimodal approach. It is important to emphasize that, while the existing evidence consistently suggests that moderate-to-vigorous intensity aerobic exercise has a significant positive impact on reducing depressive symptoms[33,58,70], this multimodal intervention has been carefully designed to provide an accessible and safe option for hospitalized patients in an acute phase of their illness. The program focuses on components that are likely to be well-tolerated by this population and, most importantly, lay the foundation for the adoption of more sustained physical activity after hospital discharge. The strategic combination of strength exercises[57] (which have shown to have a moderate effect in reducing depressive symptoms and can improve overall physical health), balance exercises[75] (to enhance stability and prevent falls), and progressive relaxation techniques[32] (to reduce muscle tension and promote relaxation), as well as a health education component, comprehensively addresses multiple dimensions of physical and mental well-being.

This protocol aligns with the growing body of evidence that supports the antidepressant effects of exercise[70] and emphasizes the importance of health education in promoting self-management and long-term treatment adherence[76]. The conceptual framework is based on the understanding of the mind-body connection and how physical activity can positively influence mood through biological mechanisms (such as the release of endorphins)[43] and psychological mechanisms (such as increased self-efficacy and improved body awareness)[77,78].

It is anticipated that the implementation of this physiotherapy program will have significant practical implications. The results could provide a solid justification for the routine inclusion of physiotherapy in treatment plans for hospitalized patients with MDD. This could translate into a tangible improvement in their perceived quality of life, a reduction in associated pain[79] (which often coexists with depression), an increase in their self-efficacy to manage their condition, and potentially greater adherence to physical activity recommendations after discharge, which in turn could help to prevent relapses[80]. On a theoretical level, this study could strengthen the evidence base for the role of physical activity and health education as effective complementary interventions in the management of MDD in an acute setting.

### Dissemination Plan

The feasibility of this protocol has been carefully assessed by designing a low-to-moderate intensity intervention that can be adapted to the individual capacities of patients and the duration of their hospital stay. Strategies have been planned to ensure the quality of the qualitative data, such as verbatim transcription of the focus groups, inductive coding performed by two researchers independently, and the triangulation of data to ensure the credibility of the findings.

The findings of this study will be disseminated through several channels to maximize their impact. Primary dissemination will occur through publication in peer-reviewed scientific journals, with PLOS ONE being the targeted journal. Presentations at national and international conferences related to physiotherapy, mental health, and healthcare will also be pursued. Furthermore, the results will be shared with participating healthcare institutions and, if applicable, to the study participants. The potential for the development of clinical practice guidelines or recommendations based on the study’s outcomes will be explored to facilitate the integration of physiotherapy into standard care for hospitalized patients with major depressive disorder.

### Study Amendments and Termination

All amendments to the study protocol will be subject to review and approval by the Aragon Ethics Committee to ensure the continued safety and ethical integrity of the research. All approved amendments will be thoroughly documented, including the date of approval, the nature of the changes, and the rationale for the modifications, and will be communicated to all relevant study personnel. The study may be terminated prematurely if unforeseen safety concerns arise, if participant recruitment is significantly below target, or if external factors impede the study’s continuation. In the event of premature termination, all collected data will be retained and analyzed to the extent possible, and the findings will be reported in accordance with ethical guidelines and applicable regulations. The reasons for termination will be clearly documented and included in any publication of the study’s results.

### Limitations

The main expected limitation is the possible loss of patients during follow-up, which may require a more extensive recruitment period to reach the target sample size. Another limitation inherent in intervention studies in the clinical setting is the difficulty of controlling the physical activity that patients may perform outside of structured sessions. The heterogeneity of major depressive disorder, with its various subtypes and clinical manifestations, could also introduce variability in responses to the intervention, although the specific diagnosis will be taken into account to minimize this risk. The lifestyle prior to admission [baseline level of physical activity] could also influence the magnitude of the changes observed.

Finally, it is crucial to recognize that patients will receive standard pharmacological and psychological treatment for their condition, which will make it difficult to exclusively attribute the observed improvements to the physiotherapy and health education interventions. However, the mixed-methods design will make it possible to explore the perceptions of patients and professionals about the specific contribution of physiotherapy to the recovery process.

## Data Availability

The feasibility of this protocol has been carefully assessed by designing a low-to-moderate intensity intervention that can be adapted to the individual capacities of patients and the duration of their hospital stay. Strategies have been planned to ensure the quality of the qualitative data, such as verbatim transcription of the focus groups, inductive coding performed by two researchers independently, and the triangulation of data to ensure the credibility of the findings. The findings of this study will be disseminated through several channels to maximize their impact. Primary dissemination will occur through publication in peer-reviewed scientific journals, with PLOS ONE being the targeted journal. Presentations at national and international conferences related to physical therapy, mental health, and healthcare will also be pursued. Furthermore, the results will be shared with participating healthcare institutions and, if applicable, to the study participants. The potential for the development of clinical practice guidelines or recommendations based on the study's outcomes will be explored to facilitate the integration of physical therapy into standard care for hospitalized patients with major depressive disorder.

## Acknowledgments

The authors wish to express their sincere gratitude to all the professional staff of the Psychiatry Department at Royo Villanova Hospital for their valuable collaboration and their great willingness to facilitate the realization of this research in their unit.

## Data Availability Statement

Deidentified research data will be made publicly available when the study is completed and published.

## Authors’ contributions

Conceptualization: J.L.P.L., C.J.S. and J.F.R.D.; Funding acquisition: J.L.P.L. and C.J.S.; Methodology: J.L.P.L., C.J.S. and R.L,U.; Project administration: C.J.S. and J.F.R.D.; Investigation: J.L.P.L., B.G.M., A.V.C. and A.P.M.; Writing—original draft preparation: J.L.P.L., C.J.S., and J.F.R.D.; Review and editing: J.L.P.L., C.J.S., R.L.U, M.G.R. and J.F.R.D.; Supervision: C.J.S., B.G.M, A.V.C. and A.P.M. All authors have read and agreed to the published version of the manuscript.

## Supporting Information

S1 Appendix. MMR-RHS checklist.

(PDF)

S2 File. Question guide.

(PDF)

S3 Table. Summary of the combined therapeutic exercise and health education intervention.

(PDF)

